# Motif-induced Subgraph Generative Learning for Explainable Neurological Disorder Detection

**DOI:** 10.1101/2024.10.27.24316244

**Authors:** Mujie Liu, Qichao Dong, Chenze Wang, Xinrui Cheng, Falih Gozi Febrinanto, Azadeh Noori Hoshyar, Feng Xia

## Abstract

The wide variation in symptoms of neurological disorders among patients necessitates uncovering individual pathologies for accurate clinical diagnosis and treatment. Current methods attempt to generalize specific biomarkers to explain individual pathology, but they often lack analysis of the underlying pathogenic mechanisms, leading to biased biomarkers and unreliable diagnoses. To address this issue, we propose a motif-induced subgraph generative learning model (MSGL), which provides multi-tiered biomarkers and facilitates explainable diagnoses of neurological disorders. MSGL uncovers underlying pathogenic mechanisms by exploring representative connectivity patterns within brain networks, offering motif-level biomarkers to tackle the challenge of clinical heterogeneity. Furthermore, it utilizes motif-induced information to generate enhanced brain network subgraphs as personalized biomarkers for identifying individual pathology. Experimental results demonstrate that MSGL outperforms baseline models. The identified biomarkers align with recent neuroscientific findings, enhancing their clinical applicability.

## 1 Introduction

Neurological disorders present a significant global health challenge, affecting millions of people worldwide [1]. The difficulty in diagnosing these disorders arises from the reliance on clinical observations rather than a deep understanding of the underlying biological mechanisms [8]. For instance, autism spectrum disorder (ASD) is diagnosed based on a spectrum of symptoms that not only vary widely among individuals but also evolve over time [2], resulting in high clinical heterogeneity. This variability underscores the need for discovering specific individual pathologies to enable accurate diagnoses and personalized treatments.

Recent advancements in functional magnetic resonance imaging (fMRI) data analysis using explainable methods have facilitated the development of biomarkers that enhance clinical diagnosis [9]. Specifically, graph neural networks (GNNs) have proven effective in extracting patterns and structures from brain activity, enabling the identification and analysis of specific subgraphs as potential biomarkers [6, 18, 24]. Some approaches identify disease-related regions based on information theory, providing biomarkers that are applicable at both group and individual levels [27, 28, 30]. However, these biomarkers often capture only superficial pathogenic differences among patients. To address this limitation, some methodologies focus on biologically significant subtypes, aiming to produce more targeted biomarkers that reflect essential pathogenic mechanisms [13, 29]. Although these methods occasionally demonstrate satisfactory performance, the biomarkers provided can be biased. This bias arises from their reliance on feature similarity analysis or clustering to generalize biomarkers across diverse symptoms without a detailed examination of the underlying pathogenic mechanisms. Such practices render these biomarkers vulnerable to imbalanced patient numbers or inconsistent subtype definitions [3]. Therefore, current methods fail to explain the clinical heterogeneity and cause biased and unreliable predictions.

To address this issue, we introduce the motif-induced subgraph generative learning (MSGL) model, a novel GNN architecture for explainable detection of neurological disorders. Recent research [7] indicates that analyzing higher-order connectivity patterns, or motifs, can provide new insights into these disorders. Inspired by this, MSGL extracts recurring connectivity patterns as motifs, serving as motif-level biomarkers to explain distinct pathologies. These motifs are integrated to guide an efficient block-wise graph generation process. By enhancing the probability distribution of connection patterns related to motifs, the resulting subgraphs effectively represent key brain connectivity structures. These subgraphs act as personalized biomarkers, facilitating more accurate and interpretable diagnostic conclusions. The main contributions of this paper are summarised as follows:

– We introduce a novel graph generative learning model, MSGL, for explainable neurological diagnoses. By delving into underlying pathogenic mechanisms, MSGL extracts motif-level biomarkers to overcome clinical heterogeneity and enhance the representational and explainable capabilities of individual pathologies.
– We developed a new recurrent-based graph generation process incorporating motif-induced information. This approach enables generated subgraphs to highlight pathology-related connectivity patterns and performs with low computational complexity.
– We conduct extensive experiments on an ASD dataset to demonstrate the effectiveness and superiority of MSGL. More importantly, visualizing the identified biomarkers reveals disease-related brain regions and abnormal connections that align with recent medical findings, showing that our method provides biological explanations.

## 2 Related Work

### 2.1 Explainable Neurological Disorder Detection

Explainable GNN-based methods are widely used to identify biomarkers and elucidate pathogenesis. For instance, BrainGNN [10] uses regions of interest (ROI)- aware graph learning to highlight important brain regions, while IBGNN [5] employs global mask learning for group and individual-level biomarkers. Recent studies suggest that edge-level explanations are more critical than node-level ones. For instance, BrainIB [30] leverages the information bottleneck principle to extract the most informative edges. Likewise, Graph-PRI [27] employs the principle of relevant information to the sparse graph structure. Additionally, CI-GNN [28] uses conditional mutual information to identify causal subgraphs that offer instance-level explanations.

However, these advancements do not fully address clinical heterogeneity and often overlook the varied pathogenesis among different symptoms. Unsupervised methods like BrainTGL [13], which uses hierarchical clustering to group patients, and BPI-GNN [29], which employs prototype learning, aim to infer diverse pathogenesis. Nevertheless, they are limited by unequal distribution of patient data across different categories or groups and inconsistent subtype definitions, leading to biased biomarkers [3]. MSGL aims to address these issues by uncovering the underlying pathogenic mechanisms of neurological disorders and facilitating explainable diagnosis.

### 2.2 Graph Generative Learning

Graph generative methods are divided into one-step and autoregressive generation methods. One-step generation models aim to create all edges between nodes in a single step. For example, variational auto-encoder-based models [14,21] infer the posterior distribution of edges to determine their probability, while generative adversarial network-based models like GraphGAN [22] use a min-max game to learn sampling from real graphs. However, generated edge probabilities in these models are independent of the latent embeddings, potentially degrading graph quality.

On the other hand, autoregressive generation builds graphs incrementally and predicts the output based on the previous step. Thus it better captures complex structural patterns. For instance, GraphRNN [26] uses two recurrent neural networks (RNNs) to generate graphs and Chu et al. [4] improved this with a random walk encoder. However, these methods are sensitive to node ordering during training. In contrast, GRAN [11] uses block sampling and an attention mechanism encoder to generate graphs effectively. Inspired by GRAN, our MSGL model incorporates motif-level biomarker information into the generation process, providing personalized biomarkers.

## 3 Methodology

The proposed framework of MSGL is shown in Figure 1. We begin with motif-level biomarker discovery through sampling and identifying brain connectivity patterns in Section 3.1. Then, in Section 3.2, we describe the motif-induced graph generative model, covering encoding, generation process, and learning objectives to identify personalized biomarkers. Finally, in Section 3.3, we explain the motif-induced subgraphs embedding method for disease diagnosis.

**Fig. 1:**
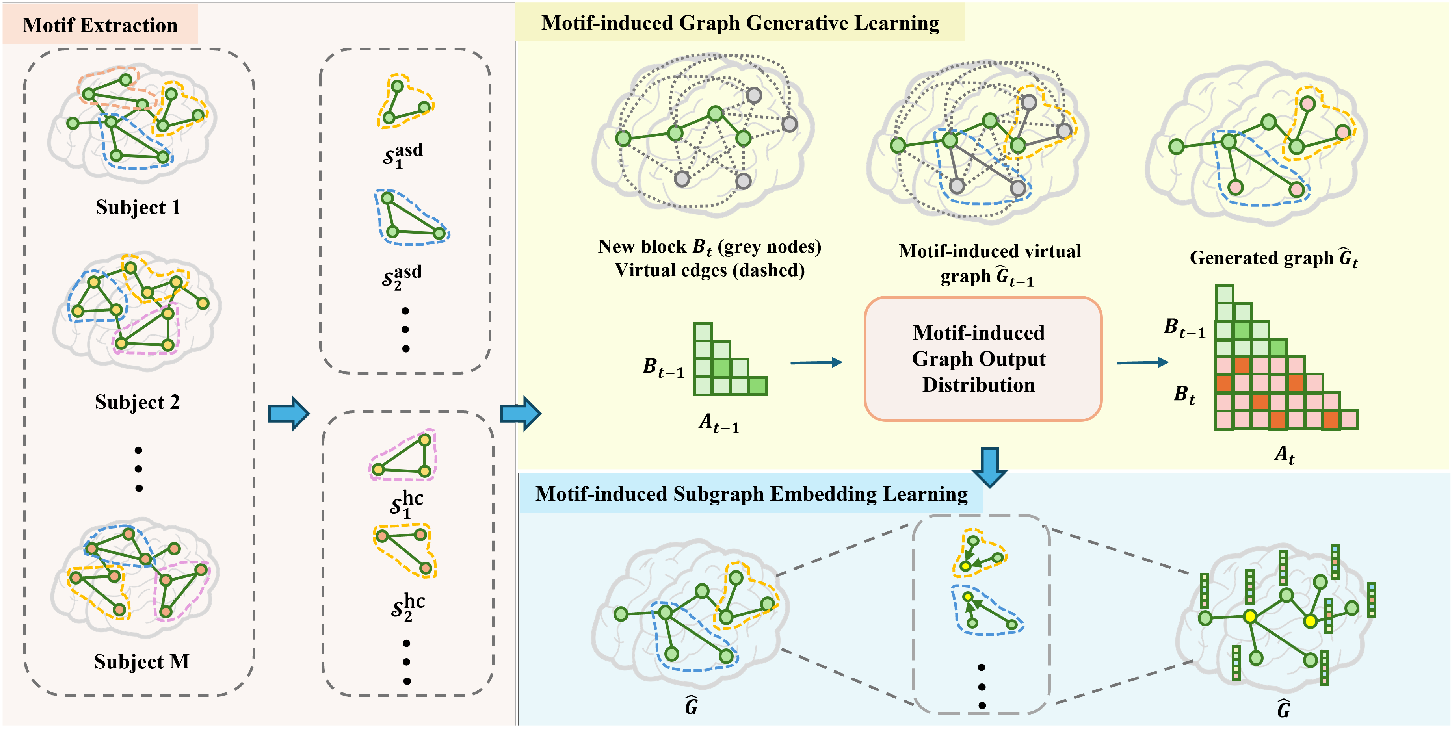
The MSGL framework is comprised of three modules: **motif extraction** to identify group-specific connection patterns, **motif-induced graph generative learning** to integrate motif information for guiding generation output, and **motif-induced subgraph embedding learning** to obtain brain representations for disorder diagnosis.

### 3.1 Motif Extraction

This module is designed to identify recurring connection patterns in brain graphs of subjects with different symptoms, classifying these patterns as motif-level biomarkers for diagnostic purposes. We consider a dataset containing brain graphs from *M* subjects, comprising both healthy controls and individuals with ASD, denoted as 𝒢 = {*G*_1_, *G*_2_, …, *G*_*M*_}. Each brain graph is defined as *G* = (*V, ε*, **A, X**), where *V* is the node set representing *N* ROIs in the brain, *ε* is the edge set, **A** ∈ ℝ^*N×N*^ is the adjacency matrix measuring the correlation strength of edges, and **X** ∈ ℝ^*N×N*^ denotes the node feature matrix, which is populated with correlation vectors. Our goal is to extract and define two distinct sets of motifs, 𝒮^hc^ and 𝒮^asd^, representing the most distinctive connection patterns for each group.

To begin, we sample connectivity structures as *S*_*t*_ = (*V*_*t*_, *ε*_*t*_) with ROIs in *V*_*t*_ and edges in *ε*_*t*_, which aims to extract a set of minimally redundant connectivity patterns covering the entire brain graph evenly: *V* = ∪_*t*_*V*_*t*_ and *ε* = ∪_*t*_*ε*_*t*_. Given the high complexity and vast scale of brain networks, direct sampling of connection patterns from the entire graph is computationally expensive. To address this challenge, we employ the farthest point sampling method [25]. This technique involves selecting nodes that are maximally distant from each other within the graph space, thus ensuring a diverse and representative sampling of connection patterns across the brain graph. Next, we identify significant recurring motifs within these graphs in the same group. To systematically evaluate the relevance and frequency of these motifs, we adopt the term frequency-inverse document frequency (TF-IDF) method [19] inspired by text mining. This technique achieves this by considering its frequency across the population relative to its occurrence within individual subjects. Using the TF-IDF values, we construct motif sets for each group as 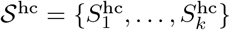 for the healthy control group and 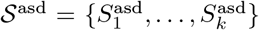 for ASD group, respectively. Ultimately, we characterize the motifs found within 𝒮^asd^ as potential motif-level biomarkers, offering a novel perspective on ASD detection.

### 3.2 Motif-induced Graph Generative Learning

This module is designed to generate deterministic subgraphs that serve as personalized biomarkers. Given an individual’s brain graph *G* and a node ordering *π*, our objective is to learn the distribution probabilities *p*(·) of potential generated nodes 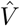 and edges 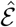 for consisting its subgraph *Ĝ*. Formally, this subgraph is generated by sequentially combining blocks 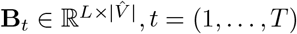, where *L* is the size of blocks and 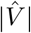 is the number of generated nodes. Therefore, the generation steps can be simplified to 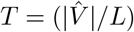 iterations. The probability of generating the graph is formulated as follows:

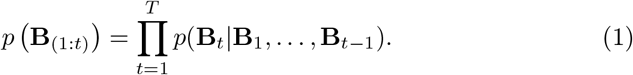

This conditional probability defines the probability of all possible edges between the nodes in the current block and the generated nodes. Once all nodes have been generated to obtain the final *p* (**B**_(1:*T*)_), the potential connection strength of edges can be formulated as **Â** = **B**_(1:*T*)_ + (**B**_(1:*T*)_)^⊺^, since brain graphs are symmetrical. Significantly, our approach generates subgraphs in a single block of rows, thereby reducing the sequence of autoregressive generation decisions by an order of magnitude 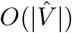, where 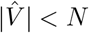. Notably, this generation process is guided by incorporating the extracted motif-level biomarkers in Section 3.1. By adjusting the generative node and edge distributions, our model ensures that the generated subgraphs retain personalized symptoms, which will be discussed in Section 3.2.

#### Graph Encoder with Attentive Messages

To accurately infer all connections of the current block in this sequential dependency generation strategy, we utilize a method of learning node generation representations by establishing virtual connections. These final node representations are then used to derive the probabilities of potential subgraphs.

In this sequential dependency generation strategy, the probability of generating a block at the current step depends on the previous steps’ probabilities, as shown in Equation (1). Thus, we denote the generated brain network subgraph at step (*t* − 1) as *Ĝ*_*t−*1_ = {**B**_1_, …, **B**_*t−*1_}, which encompass all nodes and edges from the generated blocks up to that point. Given the newly generated block **B**_*t*_ at *t*-th step, we assume that all nodes in **B**_*t*_ are connected to all other nodes, defining these connections as virtual edges. This setup allows us to formulate the potential node representations in the virtual subgraph *Ĝ*_*t*_ via a message-passing mechanism as follows:

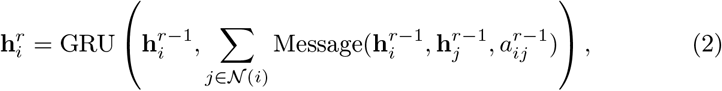

where 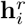 is the hidden representation for node *i* after round *r* and 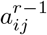 is the attention weight value of the edge between node *i* and node *j*. Specifically, the node representations in each round are aggregated using a message-passing mechanism that computes an attention-weighted sum over the neighborhood 𝒩 (*i*) of each node *i*. These representations are then updated using a gated recurrent unit (GRU). After *R* rounds, the final node representations **H**^*R*^ are obtained. Notably, we define the initial node representations as **H**^0^ = **WÂ** _*t*_ + *b*. Here, 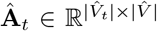 includes the connections within the virtual subgraph *Ĝ*_*t*_, where 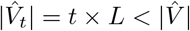, thus ungenerated parts are padded with zeros.

#### Motif-induced Graph Output Distribution

Leveraging graph encoding information, we can model the edges in the generated graph and infer blocks in the subsequent generation step. To improve their output distributions, our method integrates motif-induced information to prioritize the likelihood of generating connection patterns that typically reflect individual pathologies.

##### Generated Node Distributions

Node ordering plays a crucial role in autoregressive generative models. The number of possible random node orderings for a graph is factorial in the size of the graph, which makes exploring all potential orderings extremely complex. To mitigate this complexity and preserve the integrity of the key structural patterns within the graph, we employ a K-Core decomposition-based method for determining node orderings [16]. Typically, K-Core decomposition iteratively removes nodes with degrees less than the setting value, assigning a core number to each node, reflecting its connectivity and importance in the graph. Thus, the node ordering can be calculated within linear time |*ε*|.

To emphasize important connection patterns, we introduce motif-induced information to enhance the core number scoring of key nodes. Specifically, we apply the same sampling strategy described in Section 3.1 to the input graph *G*, matching the sampled substructures with the group-specific motifs in 𝒮^hc^ and 𝒮^asd^. We increase the degree of nodes within similar substructures to increase its K-Core ranking to obtain the final node ordering *π*. Then, the ordering will be adopted to partition the node set into several blocks in generation steps as {**B**_1_, …, **B**_*T*_} for the further edge distribution calculation.

##### Generated Edge Distributions

The probability of generating edges within the block and between the block and the already generated subgraph is determined by node representations. Specifically, using the node representations obtained after *R* rounds of message passing as described in Equation (2), we model the probability of generating edges within the current block **B**_*t*_ as a mixture of Bernoulli distributions:

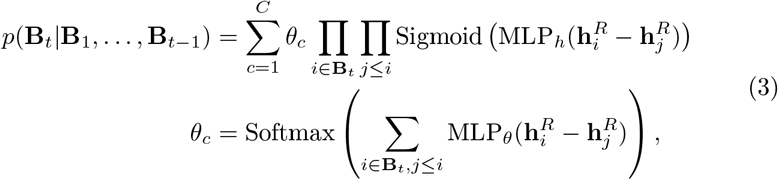

where the MLP_*h*_(·) and MLP_*θ*_(·) are two multi-layer perceptrons (MLP) networks with the *C* dimensional outputs in a mixture of Bernoulli distributions. Here, *C* is the number of mixture components that guarantee dependencies among the edges within the mixture model.To prioritize the generation of personalized connections, we incorporate motif-induced information into these distributions.

Specifically, we leverage edge centrality in the virtual graph *Ĝ*_*t*_ as mentioned in Section 3.2 to enhance the probability of generating edges 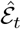 within motifs *S*_*k*_ at the *t*-th generation step. The edge centrality is calculated as the average of the connected nodes’ centrality *ϕ*(·), formalized as 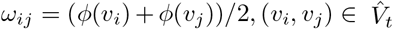. Next, we traverse the nodes in the virtual graph and match virtual substructures with group-specific motifs in 𝒮^hc^ and 𝒮^asd^. We increase the centrality of corresponding nodes to enhance their connection’s edge centrality within a similar substructure, denoted as 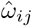. To mitigate the impact of highly densely connected nodes, we set it as log 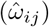. After normalizing, motif-induced edge probabilities can be formulated as:

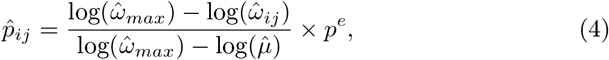

where 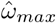 and 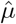 are the maximum and average of 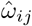 and *p*^*e*^ is a hyperparameter to control the overall probability of generating edges in the virtual graph. Finally, we multiply the calculated motif-induced edge probabilities by the probabilities calculated by the mixture of Bernoulli distributions to obtain the final graph output distribution 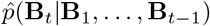.

#### Generative Learning Objective

Due to the absence of definitive criteria for our brain network subgraphs, we have designated the task of generating the entire graph as the training goal for our model. Thus, the objective function is defined as the negative log-likelihood of the variational evidence lower bound (ELBO), formulated as:

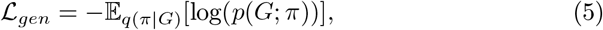

where *q*(*π*|*G*) is a variational posterior over motif-induced node ordering *π* given the graph *G*, and *p*(*G*; *π*)) is the probability distribution of the generated graph based on the given ordering.

By optimizing this formulation, our model aims to maximize the fidelity of reconstructing the input brain network graph, ensuring that the subgraphs generated during this process are biologically authentic. Finally, by controlling the step of the generation process the objective subgraph 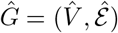, where 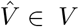 and 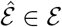 are the subset of node and edge in the original graph, is generated to represent the personalized biomarker. Overall, the detailed algorithm for our proposed methods is outlined in Algorithm 1.

##### Algorithm 1

The training algorithm.

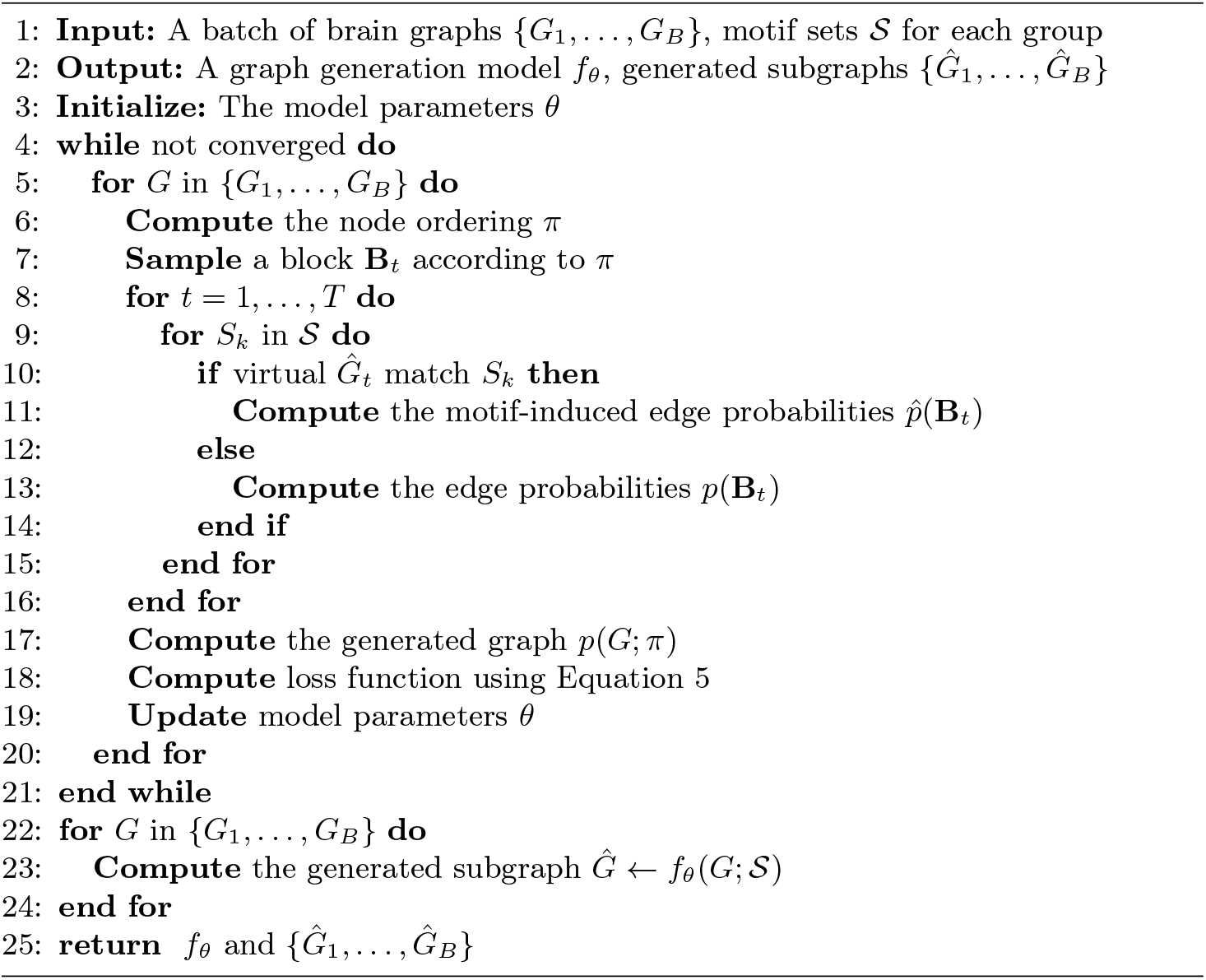

#### Computation Complexity Analysis

We thoroughly analyze the time complexity associated with the generation process of the MSGL model. We omit the batch size dimension and focus directly on the process of generating subgraphs from brain graphs. The computational complexity for the node ordering is *O*(|ε|). Subsequently, the subgraph generation process based on this node ordering has a *O*(*T*) complexity, where 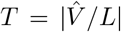. For each generation step, the computational complexity of the graph output distribution calculation is 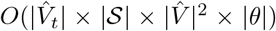, where 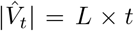 represents the number of nodes in the generated subgraph at step *t* ∈ (0, *T*), |𝒮| denotes the number of motifs, and 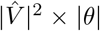 refers to the parameters of the generative network. Overall, the computational complexity can be simplified as 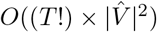.

### 3.3 Motif-induced Subgraph Representation Learning

Inspired by RH-BrainFS [25], we treat motif structures as receptive fields for disease-related ROIs. This enhances the representation learning of disease-relevant features, by aggregating the information of the central brain region, demonstrating greater brain disorder detection performance [23]. Thus, the motif-induced representation learning task can be viewed as a multi-set problem, where an MLP learns an injective function to effectively aggregate regional features within the given motifs. This process can be formalized as:

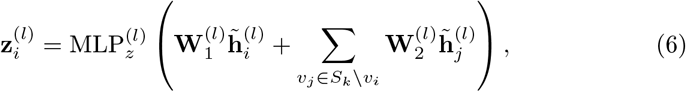

where 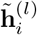is the hidden representation of central node 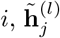 denotes the hidden representation of the other node in the motif *S*_*k*_, and 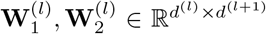 are two learnable weight matrics. Notably, the initial representation 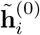is denoted as the feature vector **x**_*i*_. Finally, the obtained representation 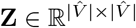 for the subgraph *Ĝ* is fed into a classifier to detect the neurological disorder.

## 4 Experiments

### 4.1 Experimental Setup

#### Datasets

This study was conducted on a real-world psychiatric dataset known as the autism brain imaging data exchange (ABIDE)^1^. ABIDE comprises restingstate fMRI data from 1,099 samples collected across 17 international sites, including individuals with ASD and healthy controls. Specifically, it consists of 528 ASD patients aged 7-64 years and 571 health control individuals aged 8.1-56.2 years, ensuring a relatively balanced class distribution. We utilize the 116 ROIs defined by the automated anatomical labeling (AAL) template and adopt Pearson correlation to construct the brain network graphs.

#### Baselines

To demonstrate the effectiveness and superiority of our MSGL model, we compared its results on the ABIDE dataset with two categories of methods. For the first category, we consider two machine learning methods, MLP and support vector machine (SVM). For the first category, four state-of-the-art explainable graph learning models specifically designed for diagnosing neurological disorders are included a benchmark method BrainGNN, and other information-theoretic methods, IBGNN, BrainIB, and CI-GNN.

#### Implementation Details

MSGL is implemented in PyTorch and experiments are conducted on an NVIDIA Tesla P100 GPU with 16GB memory. Parameters are optimized using the AdamW optimizer [15] with a learning rate of 1*e*^*−*5^. To explore pathogenic mechanisms under clinical heterogeneity, we set the number of motifs in 𝒮^hc^ and 𝒮^asd^ to 50. Balancing computational complexity and performance, we set the block size *L* of **B**_*t*_ to 5 and the number of ROIs in the generated subgraph to 45. The hyperparameter *p*^*e*^ in Equation (4) is set to 0.5.

### 4.2 Results

#### Comparison Results and Analysis

Table 1 shows the classification performance of MSGL compared to baselines on the ABIDE dataset evaluated in terms of accuracy, area under the curve (AUC), recall, and F1-score. We conducted a 5-fold cross-validation and reported the mean and standard deviation for these metrics. Extensive experiments demonstrate that MSGL outperforms all baseline models across all evaluation metrics. Specifically, MSGL achieved improvements of 2.3% in accuracy, 1.2% in AUC, 2.0% in recall, and 1.2% in F1-score compared to the best-performing baseline. The performance enhancement of MSGL can be attributed to three factors: 1) Uncovering the underlying pathogenic mechanisms of neurological disorders; 2) Motif-induced subgraph generation for a better understanding of individual pathologies; 3) A motif-induced representation learning strategy that enhances the graph’s expressiveness.

**Table 1:** The comparison results (%) on ABIDE.

| Methods | Accuracy | AUC | Recall | F1-score |
| --- | --- | --- | --- | --- |
| MLP | $60.4 \pm 7.2$ | $60.4 \pm 7.5$ | $60.4 \pm 7.2$ | $55.1 \pm 13.9$ |
| SVM | $61.4 \pm 6.5$ | $65.6 \pm 5.6$ | $61.3 \pm 6.5$ | $61.3 \pm 6.5$ |
| BrainGNN | $61.1 \pm 5.8$ | $67.5 \pm 5.1$ | $58.6 \pm 2.7$ | $66.6 \pm 4.5$ |
| IBGNN | $61.8 \pm 1.6$ | $60.7 \pm 3.4$ | $69.1 \pm 1.9$ | $68.5 \pm 2.4$ |
| CI-GNN | $67.6 \pm 1.8$ | $67.2 \pm 2.7$ | $67.6 \pm 3.9$ | <u>$72.4 \pm 2.3$</u> |
| BrainIB | $69.1 \pm 3.8$ | $69.5 \pm 1.8$ | $69.1 \pm 3.2$ | $69.0 \pm 2.1$ |
| **MSGSL (ours)** | **$71.4 \pm 2.3$** | **$70.7 \pm 1.6$** | **$71.1 \pm 2.1$** | **$73.6 \pm 1.2$** |

#### Explainable Analysis for Neurological Disorders

We validated the MSGL model using the ABIDE dataset to determine its ability to identify personalized, motif-level biomarkers for neurological disorders. We analyzed the generated sub-graphs for personalized biomarkers compared with other explainable methods.

As shown in Figure 2, the nodes are mapped onto nine brain networks with different colors, while the size of each edge reflects its weight. Our method effectively reveals enhanced connectivity between the SMN and both the LIN and FPN, aligning with previous medical research [12]. We also uncovered interactions within the LIN and SBN that might indicate unique neural patterns.

**Fig. 2:**
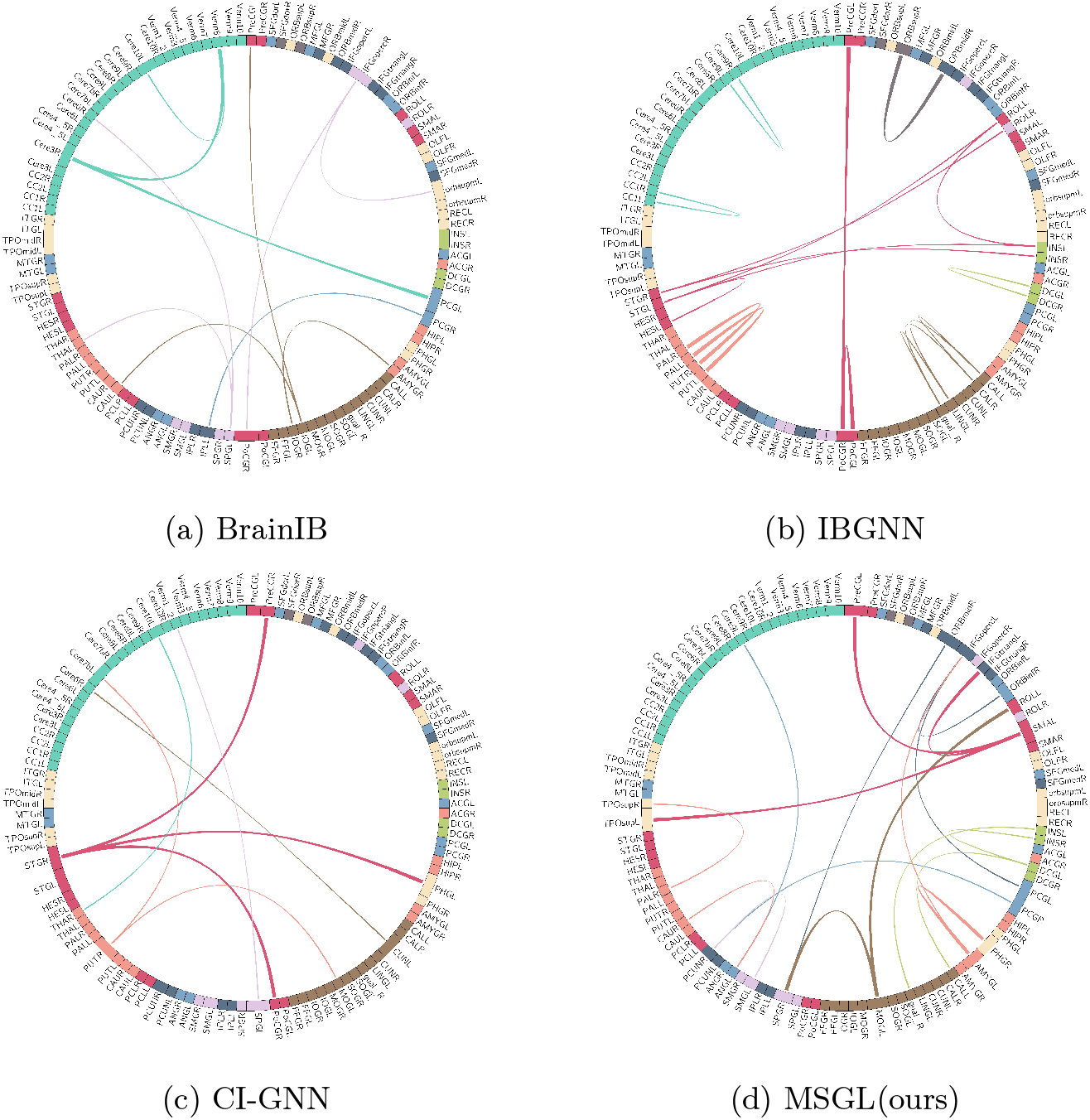
Explaination comparisons in the ASD group. The colors of brain networks are described as visual network (**VN**), somatomotor network (**SMN**), dorsal attention network (**DAN**), ventral attention network (**VAN**), limbic network (LIN), frontoparietal network (**FPN**), default mode network (**DMN**), cerebellum (**CBL**) and subcortical network (**SBN**), respectively.

Additionally, we identified four significant motif-level biomarkers, visualized in Figure 3. This visualization highlights affected regions and key connections, including the temporo-occipital areas (such as the temporal pole, middle occipital gyrus, precuneus, and right parahippocampal gyrus) and regions like the middle frontal gyrus, inferior frontal gyrus, and posterior cingulate cortex, all of which are strongly associated with ASD. These findings are consistent with previous clinical observations [17, 20].

**Fig. 3:**
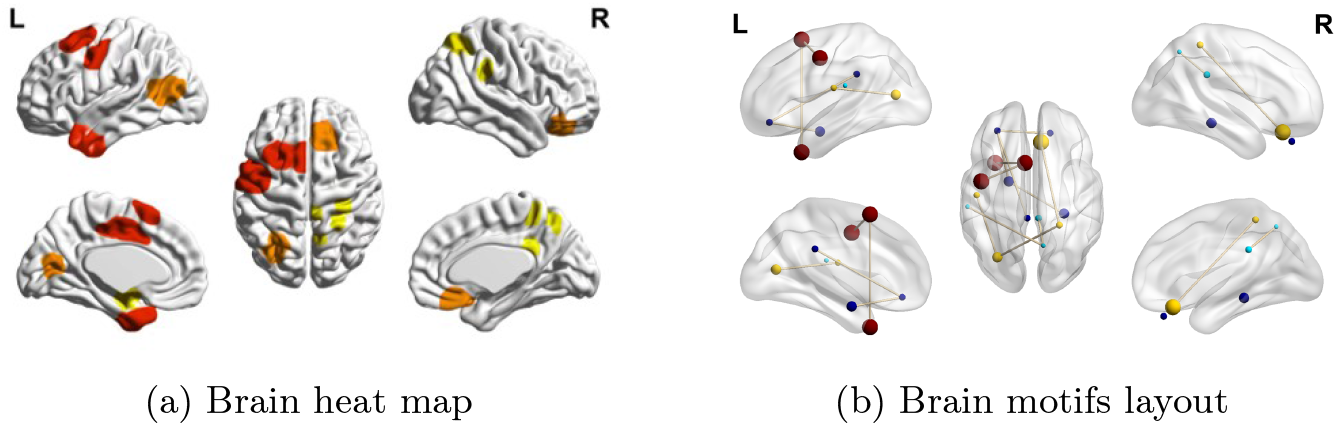
Visualization of motifs in the ASD group: Panel (a) shows shaded brain regions with darker colors indicating higher relevance. In panel (b), the color of each node denotes its motif, and the size indicates the node’s degree.

### 4.3 Ablation Study

We conducted an ablation study to assess the effects of motif-induced learning strategies on our model. We created two variants: MSGL-NoMGL, which eliminates the motif-induced graph generation strategy, relying only on traditional graph output distributions and MSGL-NoMEmb, which substitutes the motif-induced graph embedding learning with a standard neural network approach for graph learning. By comparing the classification performance of the original model with its variants, we can assess the significance of the motif-induced learning strategy. Table 2 shows that MSGL’s performance drops significantly in its variants, especially in MSGL-NoMGL, highlighting the benefit of motif-induced graph generation for identifying personalized biomarkers.

**Table 2:** The ablation study results (%) of MSGL.

| Methods | Accuracy | AUC | Recall | F1-score |
| --- | --- | --- | --- | --- |
| MSGL-NoMGL | 63.4 ± 2.1 | 62.7 ± 1.8 | 63.4 ± 2.6 | 65.2 ± 1.8 |
| MSGL-NoMEmb | 67.5 ± 3.1 | 64.8 ± 2.6 | 66.2 ± 3.4 | 67.6 ± 2.4 |
| **MSGL (ours)** | **71.4 ± 2.3** | **70.7 ± 1.6** | **71.1 ± 2.1** | **73.6 ± 1.2** |

## 5 Conclusion

In this study, we present MSGL, a novel graph generative model for explainable diagnosis of neurological disorders. MSGL identifies key motif-level biomarkers that reveal pathogenic mechanisms and enhance the explainability of individual pathologies. Our results demonstrate that MSGL surpasses other state-of-the-art methods on a real medical dataset, confirming the effectiveness of its biomarkers. Additionally, visualizations of the model results align with neurological research, providing valuable insights for future clinical diagnosis and personalized treatments.

## Data Availability

All data produced in the present study are available upon reasonable request to the authors

https://fcon_1000.projects.nitrc.org/indi/abide/

## Footnotes

1 https://fcon_1000.projects.nitrc.org/indi/abide/.

## Notes

### Competing Interest Statement

The authors have declared no competing interest.

### Funding Statement

This study did not receive any funding

### Author Declarations

https://fcon_1000.projects.nitrc.org/indi/abide/

### Summary of Updates

Superfluous image files were deleted to ensure the article's cleanliness.

